# Screen Time is Associated with Cardiometabolic and Cardiovascular Disease Risk in Childhood and Adolescence

**DOI:** 10.1101/2024.07.12.24310353

**Authors:** David Horner, Marie Jahn, Klaus Bønnelykke, Bo Chawes, Trine Flensborg-Madsen, Ann-Marie Malby Schoos, Jakob Stokholm, Morten Arendt Rasmussen

**Affiliations:** COPSAC, Copenhagen Prospective Studies on Asthma in Childhood, Herlev and Gentofte Hospital, University of Copenhagen; Unit of Medical Psychology, Department of Public Health, University of Copenhagen, 1353 Copenhagen, Denmark; Department of Pediatrics, Slagelse Hospital, Slagelse, Denmark; Section of Food Microbiology and Fermentation, Department of Food Science, University of Copenhagen

## Abstract

**Background:** Screen time in children and adolescents may be linked to cardiometabolic and cardiovascular risk. This study examines the relationship between screen time and cardiometabolic risk (CMR) factors.

**Methods:** We analysed data from over 1,000 participants in the Copenhagen Prospective Studies on Asthma in Childhood cohorts (COPSAC2010 and COPSAC2000). This longitudinal study utilised objective measures of physical activity, sleep, pubertal development, and dietary intake as covariates, and assessed mediating and moderating effects of lifestyle factors on parental- and self- and reported discretionary screen time. Our primary outcome of interest was a CMR score which was made from standardised z-scores of metabolic syndrome components (waist circumference, systolic blood pressure, HDL cholesterol, triglycerides, and glucose), adjusted for sex and age. Secondary outcomes were insulin resistance, inflammation, atherogenic lipoproteins, and anthropometric measures. We utilised supervised machine learning modelling of blood NMR metabolomics to identify a unique metabolic signature of screen time. Finally, we assess screen time associations with a predicted Cardiovascular Risk Score derived from Cox proportional hazards models of 10-year CVD events trained in the UK Biobank.

**Results:** Increased screen time was significantly associated with CMR in children and adolescents, with each additional hour of screen time linked to a higher CMR z-score (children at 10-years: 0.08 [0.01 - 0.14], p=0.021; adolescents at 18-years: 0.13 [0.07 - 0.2], p=0.001). In childhood, sleep duration (p=0.029) and time of onset (p=0.009) significantly moderated the relationship between screen time and CMR; less sleep combined with high screen time significantly increased cardiometabolic risk. In adolescence, sleep duration likewise significantly moderated the association between screen time and CMR (p=0.012), replicating the findings from childhood. A supervised machine learning model trained in the childhood cohort identified a unique metabolic signature in the blood NMR metabolome associated with screen time, which was validated in the adolescent cohort (0.14 [0.03-0.26], p=0.014). CVD-risk scores modelled from CVD-events were directionally associated with screen time in childhood (0.06 [−0.02 - 0.13], p=0.15) and significantly associated with screen time in adolescence (0.07 [0.01 - 0.13], p=0.017) in fully adjusted models.

**Conclusion:** Increased screen time is significantly associated with higher cardiometabolic risk in children and adolescents, with sleep duration moderating this relationship. A unique metabolic signature of screen time was validated across cohorts, and screen time was associated with higher cardiovascular risk scores in adolescence. These findings underscore the importance of considering screen time and sleep duration in addressing cardiometabolic and cardiovascular risks.

## INTRODUCTION

Cardiovascular disease (CVD) is a leading cause of morbidity and mortality worldwide and has its roots in childhood (1). Key predictors of early onset of CVD are the presence of cardiometabolic risk (CMR) factors (2), such as components of the metabolic syndrome, insulin resistance, inflammation, ApoB-containing lipoproteins and obesity (3,4).

As we transition into the digital age, these risks may be exacerbated as children and adolescents are spending an increasing amount of time engaged with screens and digital content, be it for educational, recreational, or social purposes. Emerging research has begun to shed light on the potential health implications of this trend, with several studies noting associations between screen time and CMR factors in childhood and adolescence (5–7). However, the relationship between screen time and CMR is complex and likely multifactorial. Reductions in sleep time (8), increased sedentary time (9), reduced physical activity (10), and unhealthy dietary patterns (11) have all been associated with increased screen time. It is plausible that they may serve as moderators, or mediators, in the relationship between screen time and CMR (12). This interplay is important to investigate as prospective studies show that increased screen time in adolescence is associated with a higher risk of obesity, elevated waist circumference, and diabetes in adulthood (6). Previous studies investigating this relationship have lacked objective measures for contextual lifestyle factors, relying on subjective self-reported measures of diet, sleep, and physical activity, and thus may be subject to bias and inaccuracies (13).

In this study, we aim to address these limitations by assessing longitudinal associations between screen time and CMR factors in two mother-child cohorts. By juxtaposing the two cohorts from the Copenhagen Prospective Studies on Asthma in Childhood (COPSAC), we gain valuable insights into how screen time patterns and their potential impact on CMR evolve from childhood through adolescence. We hypothesise that higher screen time during childhood and adolescence is associated with adverse CMR, as defined by the components of the metabolic syndrome. By using targeted blood metabolomics, we identify a metabolic signature of screen time, showing that screen time-associated metabolic disturbances are robust predictors across cohorts. Finally, we link screen time to CVD outcomes using an NMR CVD score trained in a large prospectively followed adult cohort.

## METHODS

### Study Design

The Copenhagen Prospective Studies on Asthma in Childhood 2010 mother-child cohort (COPSAC2010) is a prospective general population study that consists of 700 mother-child pairs with extensive phenotyping from 14 clinical visits and exposure assessments since birth, up to the age of 10 years (14). COPSAC2000 is likewise a prospective mother-child cohort consisting of 411 children born of asthmatic mothers with extensive phenotyping from 19 clinical visits and exposure assessments since birth, up to the age of 18 years (15). Participants in COPSAC2010 (n=2) and COPSAC2000 (n=1) with type 1 diabetes were excluded from the analysis due to potential confounding (16).

### Screen time Measurement

Screen time, the primary exposure, was measured using questionnaire responses from the COPSAC2010 and COPSAC2000 cohorts, focusing on discretionary screen time. In COPSAC2010, parents reported children’s average screen time at 6 and 10 years on weekdays and weekends. In COPSAC2000, 18-year-olds detailed screen time from Monday to Thursday and Friday to Sunday, differentiated by type. Total screen time was calculated and weighted averages were derived. *Further details are in the Supplementary Methods*.

### Cardiometabolic Outcomes

Our primary outcome was a CMR score derived from the components of the metabolic syndrome (17,18). Prior to calculating CMR scores at each clinical visit, the five measures (waist circumference, systolic blood pressure (SBP), high-density lipoprotein (HDL) cholesterol, triglycerides and glucose were adjusted for sex and age, SBP was further adjusted for height (19). The total CMR score was then calculated by adding internally (within each cohort) standardised z-scores of waist circumference, SBP, negative HDL cholesterol, logged triglycerides, and glucose, and then dividing the sum by the square-root of 5 (17,18).

Secondary outcomes include other established CMR risk factors including HbA1C, Homeostatic Model Assessment for Insulin Resistance (HOMA-IR), high sensitivity CRP (hs-CRP), GlycA, Apolipoprotein B (ApoB) and numerous measurements of body anthropometrics. Furthermore, we included an NMR Cardiovascular Risk Score based on sex-stratified Cox proportional hazards models for 10-year CVD risk trained in the UK Biobank (20). Our analysis utilised the sex-stratified model coefficients from this work; by internally z-scoring the individual model components in the COPSAC cohorts, multiplying them by the model coefficients, and summing these components into a total CVD-risk score. We subsequently z-scored for each of the respective COPSAC cohorts, to facilitate interpretation of estimates. Nightingale targeted Blood NMR data was available at 10 years for COPSAC2010 and 18 years for COPSAC2000. *Further details are in the Supplementary Methods*.

### Body Anthropometrics

Anthropometrics were assessed at each clinical visit. Body composition analysis at age 10 in COPSAC2010 and age 18 in COPSAC2000 was measured by Bioelectrical Impedance Analysis (BIA) using a Tanita scale (Health monitor, version 3.2.7) to derive muscle mass (kg), fat mass (kg) and bone mass (kg). Fat free mass (FFM) (body weight - fat mass), fat mass index (fat mass / (height²)) and fat free mass index (fat free mass/ (height² (m))) were derived from these measurements. *Further details are in the Supplementary Methods*.

### Accelerometer-derived sleep and activity data

Activity and sleep information was derived from accelerometry data (Actigraph GT3X+, 30Hz) over 14 days using the GGIR package in R (version 2.9.0) (21–24). *Further details are in the Supplementary Methods*.

### Covariates

Analysis was conducted unadjusted and fully adjusted. Unadjusted analysis covariates included only sex and age, as there are well documented sex and age-dependent effects on our outcome measures (25,26). Fully adjusted multivariable analysis in both cohorts were further adjusted for social circumstances (principal component 1 of a principal component analysis (PCA) of household income, maternal education level, and maternal age), maternal smoking during pregnancy, number of siblings, and accelerometer derived sedentary time, symptoms of attention-deficit hyperactivity disorder (ADHD), light activity time, moderate to vigorous activity time (a sum of moderate and vigorous activity time), sleep duration, time of sleep onset. In COPSAC2010, we included further adjustment for objective markers of pubertal progression using the gonadotropic hormones, luteinizing hormone (LH) and follicle-stimulating hormone (FSH), which increase at puberty onset (27). LH and FSH were included in models as an interaction effect with child sex, to account for potential differences between sexes. Additionally, in COPSAC2010, adjustments were made for dietary patterns assessed at the age of 10 years. *Further details are in the Supplementary Methods*.

### Statistical Analysis

Linear regression models were used to assess the associations of screen time on CMR factors and anthropometric outcomes. As we had repeated measurements of outcome data at 6 and 10 years in COPSAC2010, we utilised linear mixed models from the lme4 R package (version ‘1.1.28’) with a fixed slope and variable intercept for our primary outcome analyses (28). All interaction models and mediation models were multivariable. The mediation package in R was used for mediation analysis (version ‘4.5.0’) (29). In our models for CMR outcomes, we apriori decided not to include child’s anthropometrics as covariates. This was based on the potential for anthropometrics to act as a causal intermediary between screen time and CMR outcomes.

Gaussian graphical models were utilised to illustrate the non-zero relationships (95% Cl) between screen time, covariates, and total CMR, controlling for the linear effects of all covariates expressed as partial correlations.

To further explore the relationship between screen time and its metabolic associations between cohorts, we employed supervised machine learning modelling - sparse partial least squares (sPLS). The sPLS model was trained in COPSAC2010 using the Nightingale Health Ltd high-throughput targeted NMR-metabolomics platform as the predictor and screen time as the outcome variable (30). Screen time was z-scored relative to each cohort for this analysis, allowing comparison of estimates.

A two-sided P < 0.05 was considered statistically significant. All statistical analyses were carried out using R (version 4.1.1). *Further details are in the Supplementary Methods*.

## RESULTS

### Baseline Characteristics

For the COPSAC2010 cohort, screen time was available for 657 children (94.1%) at 6 years, and for 630 children (90.3%) at 10 years. For COPSAC2000, screen time was available for 364 adolescents (88.6%). Marked differences in screen time were observed between the cohorts. Average screen time in COPSAC2010 was 2.0 hours (SD=0.9) at 6 years, and 3.2 hours (SD=1.2) at 10 years, representing a significant increase over time (p<0.001). COPSAC2000 at 18 years had a significantly higher average screen time of 6.1 hours (SD=2.1) (p<0.001) (*Figure S1*). Baseline characteristic differences between the cohorts included parental income, maternal education, maternal age at birth, gestational age, maternal smoking in pregnancy, and number of siblings (p<0.001) (*Table S1*).

In COPSAC2010, screen time was positively associated with age, male sex, sedentary time, sleep onset, ADHD symptoms, and a Western dietary pattern, and negatively associated with light and moderate activity time and sleep duration (*Table S2, dietary patterns illustrated in Figure S2*). Similarly, in COPSAC2000 screen time had significant positive associations with male sex and maternal smoking during pregnancy, and negatively associated with social circumstances, and sleep duration (*Table S2*). Moreover, with respect to sex differences, males in COPSAC2010 used more screen time at 10 years compared to females (3.4 vs. 3.0, p<0.001), whereas no significant difference was noted at 6 years (p=0.196). In COPSAC2000, a similar sex difference in screen time was noted (6.6 vs 5.7, p<0.001). Both cohorts demonstrated numerous significant sex-related differences across CMR profiles, anthropometrics, and covariates (*Table S3*).

Between cohorts, differences in covariates such as age, maternal smoking, sedentary time, physical activity, and sleep patterns were identified (p<0.001). All CMR factors were significantly different between cohorts (p<0.001), except for ApoB. We computed a total CMR score by summing z-scores of waist circumference, SBP, HDL cholesterol, triglycerides, and glucose levels (18) in both cohorts. Cohort differences were noted for all measures of body anthropometrics (p<0.001) (*Table 1*). Within cohorts, correlations between screen time and model covariates (*Figure S3*), and correlations between CMR factors (*Figure S4)* are further visualised in comprehensive heatmaps.

**Table 1.** Exposure, Outcome and Covariate Data for COPSAC2010 and COPSAC2000. This table presents the baseline characteristics, outcomes, and covariates for the COPSAC2010 (n=630) and COPSAC2000 (n=364) cohorts. It includes data on screen time, cardiometabolic risk factors, anthropometrics, and lifestyle behaviours such as physical activity and sleep patterns. Significant differences between the cohorts are indicated, including differences in screen time, CMR factors, and anthropometrics. The data presented in this table provide a comprehensive overview of the cohorts’ characteristics. * Glucose is estimated from HBA1C in COPSAC2010.

| COPSAC2010 and COPSAC2000 Comparison | COPSAC2010 | COPSAC2000 | p | C2010/C2000<br>% Missing |
| --- | --- | --- | --- | --- |
| n = | 630 | 364 | - | - |
| **Screen Time and Cardiometabolic Outcomes** | - | - | - | - |
| Average Screen Time (hours) (mean (SD)) | 3.20 (1.21) | 6.11 (2.11) | <0.001 | 0 / 0 |
| Waist Size (cm) (mean (SD)) | 64.75 (7.42) | 80.50 (11.34) | <0.001 | 4.8 / 1.1 |
| Systolic Blood Pressure (mean (SD)) | 103.94 (6.93) | 115.78 (9.99) | <0.001 | 5.6 / 1.4 |
| HDL mmol/l (mean (SD)) | 1.56 (0.32) | 1.22 (0.26) | <0.001 | 20.0 / 8.0 |
| Triglyceride mmol/l (mean (SD)) | 0.74 (0.32) | 0.97 (0.44) | <0.001 | 19.8 / 8.0 |
| Glucose (mmol/l) (mean (SD)) | 4.18 (0.36) | 5.10 (0.40) | <0.001 | 16.8 / 8.0 |
| HB1AC (mmol/mol) (mean (SD)) | 32.10 (2.68) | 31.36 (2.80) | <0.001 | 19.2 / 8.8 |
| HOMA-IR (mean (SD)) | 1.96 (0.25) | 2.78 (1.92) | <0.001 | 20.8 / 9.3 |
| High sensitivity CRP (mg/L) (mean (SD)) | 0.57 (1.54) | 1.71 (2.55) | <0.001 | 46.3 / 10.2 |
| GlycA (mmol/l) (mean (SD)) | 0.68 (0.07) | 0.76 (0.11) | <0.001 | 16.8 / 17.9 |
| ApoB (g/l) (mean (SD)) | 0.69 (0.12) | 0.68 (0.15) | 0.207 | 16.8 / 17.9 |
| **Cohorts Characteristics and Covariates** | - | - | - | - |
| Male Sex (%) | 324 ( 51.4) | 179 (49.2) | 0.536 | 0 |
| Age (mean (SD)) | 10.30 (0.39) | 17.73 (0.57) | <0.001 | 0 |
| Siblings (mean (SD)) | 1.47 (0.94) | 1.24 (0.87) | <0.001 | 0 |
| Maternal Pregnancy Smoking (%) | 45 ( 7.1) | 83 (22.8) | <0.001 | 0 |
| Sedentary time (hours) (mean (SD)) | 8.47 (1.20) | 10.91 (1.31) | <0.001 | 23.8 / 22.8 |
| Light activity time (hours) (mean (SD)) | 3.84 (0.66) | 3.52 (0.75) | <0.001 | 23.8 / 22.8 |
| Moderate Activity time (hours) (mean (SD)) | 2.15 (0.53) | 1.59 (0.55) | <0.001 | 23.8 / 22.8 |
| Vigorous activity time (hours) (mean(SD)) | 0.42 (0.21) | 0.09 (0.10) | <0.001 | 23.8 / 22.8 |
| Sleep (mean (SD)) | 9.04 (0.76) | 7.73 (0.86) | <0.001 | 23.8 / 22.8 |
| Sleep Onset Time (hours) (mean (SD)) | 21.84 (0.82) | 24.46 (1.19) | <0.001 | 23.8 / 22.8 |
| N of valid days (Accelerometer) (mean (SD)) | 12.32 (1.66) | 11.70 (2.13) | <0.001 | 23.8 / 22.8 |
| **Body Anthropometric** | - | - | - | - |
| BMI (mean (SD)) | 17.08 (2.38) | 22.91 (4.10) | <0.001 | 11.1 / 2.7 |
| Body Weight (kilograms) (mean (SD)) | 35.77 (6.98) | 70.14 (14.06) | <0.001 | 10.8 / 2.2 |
| Fat Mass (kilograms) (mean (SD)) | 7.99 (3.24) | 16.97 (8.28) | <0.001 | 11.1 / 2.7 |
| Fat percent (%) (mean (SD)) | 21.71 (4.78) | 23.70 (8.22) | <0.001 | 11.1 / 2.7 |
| Muscle Mass (kilograms) (mean (SD)) | 26.31 (4.10) | 50.46 (9.71) | <0.001 | 11.1 / 2.7 |
| Skeletal Mass (kilograms) (mean (SD)) | 15.73 (2.44) | 30.26 (5.93) | <0.001 | 11.1 / 2.7 |
| Bone Mass (kilograms) (mean (SD)) | 1.48 (0.22) | 2.68 (0.48) | <0.001 | 11.1 / 2.7 |
| Fat Free Mass (kilograms) (mean (SD)) | 27.79 (4.31) | 53.14 (10.19) | <0.001 | 11.1 / 2.7 |
| Fat Mass Index (FMI) (mean (SD)) | 3.80 (1.38) | 5.66 (2.92) | <0.001 | 11.3 / 2.7 |
| Fat-free Mass Index (FFMI) (mean (SD)) | 13.28 (1.22) | 17.28 (2.14) | <0.001 | 11.3 / 2.7 |

### Screen Time is associated with Cardiometabolic Risk in COPSAC2010

In COPSAC2010, we used mixed models to assess the association between screen time and total CMR, and its constituent components, at 6 and 10 years. After adjusting for relevant confounders, there was a significant positive association between screen time (per hour increase) and CMR (0.08 [0.01 - 0.14], p=0.021) (*Table 2*). In males, the association for CMR (0.10 [0.02 - 0.19], p=0.013) was directionally stronger than in females (0.02 [−0.08 - 0.12], p=0.694), but there was no significant interaction between sexes (*Table S4*). Cross-sectionally at 10 years, screen time associations with CMR were stronger than at 6 years (0.16 [0.05 - 0.27], p=0.007 vs. 0.06 [−0.06 - 0.17], p=0.321) (*Table 2, Table S5*). Further adjustment for the COPSAC2010 prenatal interventions with n3-LCPUFA and high-dose vitamin D did not alter these associations (*Table S6*).

**Table 2.** Associations between Screen Time and Cardiometabolic Risk Factors in COPSAC2010 and COPSAC2000. Results of mixed models and linear regression analyses assessing the associations between screen time and total CMR, as well as its individual components, markers of insulin resistance, inflammation and atherogenic lipoproteins in the COPSAC2010 and COPSAC2000 cohorts. The table provides both unadjusted and adjusted associations, with the latter controlling for potential confounders (social circumstances, maternal smoking during pregnancy, number of siblings, sedentary time, light activity time, moderate to vigorous activity time, sleep duration and time of sleep onset. In COPSAC2010, we included further adjustment for luteinizing hormone (LH) and follicle-stimulating hormone (FSH). The associations are presented as estimates with 95% confidence intervals and corresponding p-value. † Glucose was not available at 6 years in COPSAC2010, and thus substituted with HBA1C for the purposes of mixed modelling.

| **COPSAC2010 6 & 10 year Mixed Model** | **n =** | **Unadjusted [95% CI] p-value** | **Adjusted [95% CI] p-value** |
| --- | --- | --- | --- |
| Cardiometabolic Risk (Z-score) | 513/525 | 0.08 [0.02 - 0.15] (p = 0.009) | 0.08 [0.01 - 0.14] (p = 0.021) |
| Waist size (cm) | 621/600 | 0.22 [-0.08 - 0.52] (p = 0.157) | 0.2 [-0.11 - 0.5] (p = 0.21) |
| Systolic Blood Pressure (mmHg) | 605/595 | 0.09 [-0.27 - 0.45] (p = 0.621) | 0.06 [-0.31 - 0.42] (p = 0.758) |
| HDL Cholesterol (mmol/l) | 491/504 | -0.02 [-0.04 - 0] (p = 0.027) | -0.02 [-0.03 - 0] (p = 0.052) |
| Triglycerides (mmol/l)** Logged | 465/505 | 0.03 [0.01 - 0.06] (p = 0.006) | 0.03 [0.01 - 0.06] (p = 0.013) |
| Glucose (mmol/l) † | 507/509 | 0 [-0.03 - 0.03] (p = 0.998) | 0 [-0.03 - 0.03] (p = 0.908) |
| **COPSAC2010 10 year Linear Model** | **n=** | **Unadjusted [95% CI] p-value** | **Adjusted [95% CI] p-value** |
| Cardiometabolic Risk (Z-score) | 525 | 0.14 [0.06 - 0.22] (p = 0.001) | 0.12 [0.03 - 0.2] (p = 0.011) |
| Waist size (cm) | 600 | 0.47 [-0.03 - 0.98] (p = 0.068) | 0.34 [-0.19 - 0.88] (p = 0.211) |
| Systolic Blood Pressure (mmHg) | 595 | 0.19 [-0.29 - 0.66] (p = 0.446) | 0.19 [-0.32 - 0.7] (p = 0.458) |
| HDL Cholesterol (mmol/l) | 504 | -0.03 [-0.05 - 0] (p = 0.037) | -0.02 [-0.05 - 0] (p = 0.091) |
| Triglycerides (mmol/l)** Logged | 505 | 0.06 [0.03 - 0.08] (p <0.001) | 0.05 [0.02 - 0.07] (p = 0.003) |
| Glucose (mmol/l) | 524 | 0.01 [-0.02 - 0.04] (p = 0.494) | 0.01 [-0.02 - 0.04] (p = 0.467) |
| HB1AC (mmol/mol) | 509 | 0.01 [-0.19 - 0.21] (p = 0.928) | 0 [-0.21 - 0.21] (p = 0.987) |
| HOMA-IR | 499 | 0.02 [0 - 0.04] (p = 0.013) | 0.02 [0 - 0.04] (p = 0.024) |
| High sensitivity CRP (mg/L) | 338 | 0.03 [-0.12 - 0.17] (p = 0.724) | -0.02 [-0.17 - 0.13] (p = 0.827) |
| GlycA (mmol/l) | 524 | 0.01 [0 - 0.01] (p = 0.01) | 0.01 [0 - 0.01] (p = 0.053) |
| ApoB (g/l) | 524 | 0 [-0.01 - 0.01] (p = 0.927) | 0 [-0.01 - 0.01] (p = 0.961) |
| NMR Cardiovascular Risk Score (Z-score) | 520 | 0.07 [0 - 0.14] (p = 0.054) | 0.06 [-0.02 - 0.13] (p = 0.15) |
| **COPSAC2000 18 year Linear Model** | **n=** | **Unadjusted [95% CI] p-value** | **Adjusted [95% CI] p-value** |
| Cardiometabolic Risk (Z-score) | 335 | 0.14 [0.08 - 0.2] (p <0.001) | 0.13 [0.07 - 0.2] (p <0.001) |
| Waist size (cm) | 360 | 1.47 [0.93 - 2.01] (p <0.001) | 1.3 [0.76 - 1.84] (p <0.001) |
| Systolic Blood Pressure (mmHg) | 359 | 0.7 [0.23 - 1.16] (p = 0.004) | 0.63 [0.15 - 1.11] (p = 0.011) |
| HDL Cholesterol (mmol/l) | 335 | -0.01 [-0.03 - 0] (p = 0.06) | -0.01 [-0.03 - 0] (p = 0.032) |
| Triglycerides (mmol/l)** Logged | 335 | 0.02 [0 - 0.04] (p = 0.042) | 0.02 [0 - 0.04] (p = 0.103) |
| Glucose (mmol/l) | 335 | 0 [-0.02 - 0.02] (p = 0.675) | 0.01 [-0.02 - 0.03] (p = 0.611) |
| HB1AC (mmol/mol) | 333 | -0.08 [-0.23 - 0.07] (p = 0.282) | -0.09 [-0.24 - 0.06] (p = 0.24) |
| HOMA-IR | 330 | 0.09 [-0.01 - 0.19] (p = 0.064) | 0.08 [-0.02 - 0.18] (p = 0.13) |
| High sensitivity CRP (mg/L) | 327 | 0.04 [-0.09 - 0.17] (p = 0.541) | 0.01 [-0.13 - 0.15] (p = 0.863) |
| GlycA (mmol/l) | 299 | 0.01 [0.01 - 0.02] (p <0.001) | 0.01 [0.01 - 0.02] (p <0.001) |
| ApoB (g/l) | 299 | 0.01 [0.01 - 0.02] (p = 0.001) | 0.01 [0.01 - 0.02] (p = 0.001) |
| NMR Cardiovascular Risk Score (Z-score) | 299 | 0.08 [0.02 - 0.13] (p = 0.007) | 0.07 [0.01 - 0.13] (p = 0.017) |

To illustrate the relationship between CMR and covariates in the COPSAC2010 cohort, we utilised gaussian graphical models (*Figure 1A*, sex stratified models *Figure S5).* Figure 1A highlights the significant partial correlations between screen time and CMR in COPSAC2010, even when accounting for model covariates. These models revealed an association between time of sleep onset and CMR and given the highly co-linear relationship between sleep duration and onset, we assessed if either may act as a potential mediator in the association between screen time and CMR. A mediation analysis indicated that 12.0% (p=0.030) of the association between screen time and CMR was mediated through sleep duration, and not by time of sleep onset (p=0.7).

**Figure 1.**
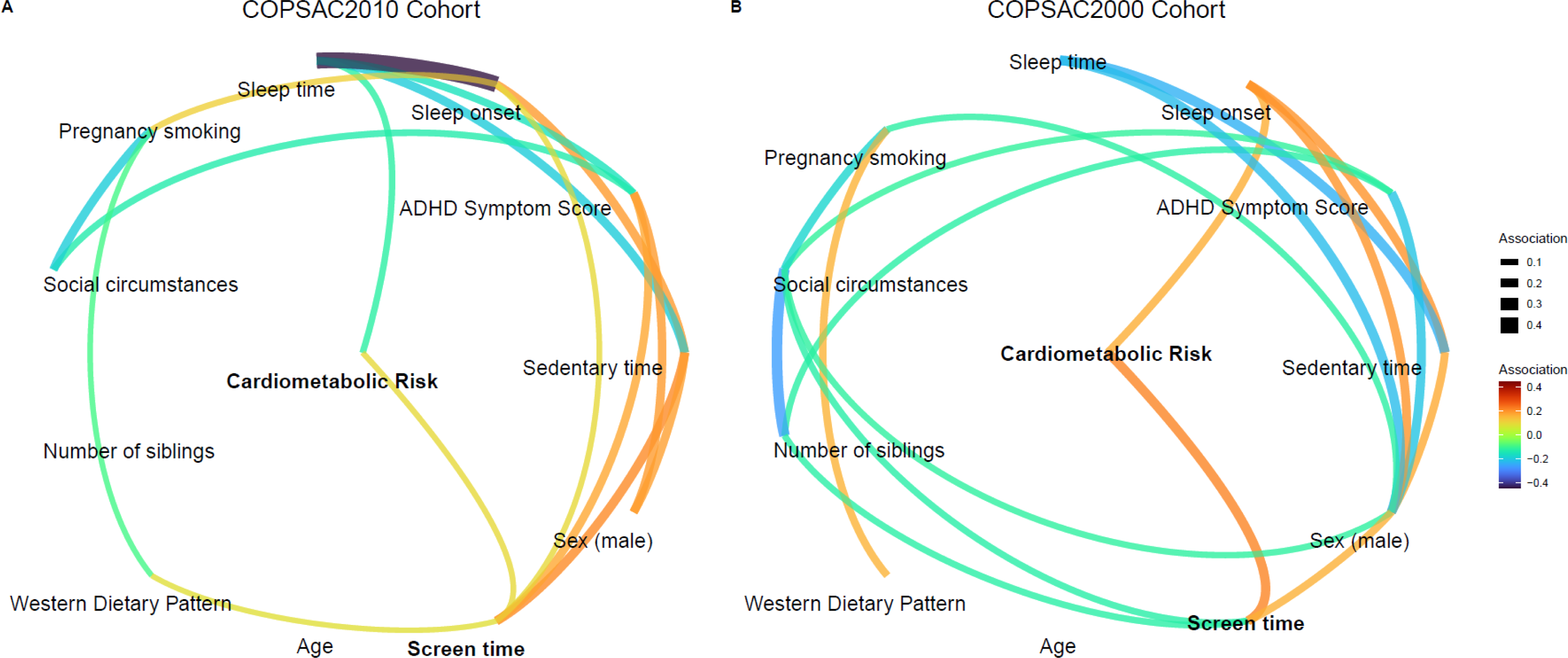
Gaussian Graphical Models Depicting the Integrated Relationships between Cardiometabolic Risk, Screen Time, and Other Covariates in COPSAC2010 and COPSAC2000. Graphical models illustrate relationships (95% Cl) between screen time,cardiometabolic risk and model covariates. Panel A represents the COPSAC2010 cohort, showing that screen time and a Western dietary pattern at 6 years are independently associated with cardiometabolic risk, when accounting for model covariates. Panel B represents the COPSAC2000 cohort, indicating that both screen time and sleep onset have independent associations with cardiometabolic risk.

Using screen time data available from 6 years, we conducted a series of sub-analyses to provide longitudinal insights into the relationship between screen time and CMR factors at 10 years. An increase per hour of screen time from 6 to 10 years was associated with measures of insulin resistance, with an increase in HOMA-IR (0.03 [0.01 - 0.04], p=0.006). Moreover, screen time at 10 years, adjusted for screen time at 6 years, was significantly associated with total CMR (0.11 [0.02 - 0.2], p=0.020) (*Table S5*). Associations stratified by weekday and weekend screen time can be seen in Table S7.

### Screen Time is associated with Cardiometabolic Risk in COPSAC2000

In COPSAC2000 at age 18 years, we likewise identified significant positive associations between screen time and CMR (0.13 [0.07 - 0.2], p<0.001). Furthermore, several CMR factors were significantly associated including waist circumference (1.30 [0.76 - 1.84], p<0.001), SBP (0.63 [0.15 - 1.11], p = 0.011), HDL cholesterol (−0.01 [−0.03 - 0], p=0.032), GlycA (0.01 [0.01 - 0.02], p<0.001) and ApoB (0.01 [0.01-0.02], p=0.001) (*Table 2*). Sex-stratified analysis again revealed stronger associations for males, for CMR (0.14 [0.06-0.22], p=0.001), waist circumference (1.56 [0.89 - 2.22], p<0.001), and SBP (0.9 [0.24 - 1.56], p=0.008). However, no statistically significant interaction effects were observed between sexes. Further adjustments for a Western dietary pattern metabolome score, derived from newborn dry blood spots, did not meaningfully change these findings (*Table S6*).

To illustrate the relationship between CMR and covariates in the COPSAC2000 cohort, we utilised gaussian graphical models (*Figure 1B*, sex stratified models *Figure S5)*. Figure 1B highlights the robust association between screen time and total CMR in COPSAC2000, when accounting for model covariates. Sleep onset time also had an independent association with CMR; however, there was no association between sleep onset and screen time, suggesting an independent association.

Screen time, whether through phones, TVs, or gaming, consistently demonstrated a positive association with multiple CMR factors (*Table S8*). Associations stratified by weekday and weekend screen time can be seen in Table S7.

### Screen Time and Body Anthropometry in COPSAC2010 and COPSAC2000

In COPSAC2010, at age 10 years, we found no significant associations between screen time and various body anthropometric measures, including BMI, body weight, fat mass, fat percent, muscle mass, skeletal mass, bone mass, fat-free mass, fat mass index (FMI), and fat-free mass index (FFMI) in both females and males (*Table 3*).

**Table 3.** Sex-Stratified Associations between Screen Time and Body Anthropometrics in COPSAC2010 and COPSAC2000. Results of adjusted sex-stratified analyses assessing the associations between screen time and various body anthropometric measures in the COPSAC2010 and COPSAC2000 cohorts. The measures include BMI, body weight, fat mass, fat percent, muscle mass, skeletal mass, bone mass, fat-free mass, fat mass index, and fat-free mass index. The associations are presented separately for females and males in each cohort. The results highlight the significant associations between increased screen time and various body anthropometric measures, with differences noted between females and males. The associations are presented as estimates with 95% confidence intervals and corresponding p-values.

| **Body Anthropometrics COPSAC2010** | **n=** | **Female Adjusted [95% CI] p-value** | **n=** | **Male Adjusted [95% CI] p-value** | **P-Interaction** |
| --- | --- | --- | --- | --- | --- |
| BMI (actual) | 269 | 0.11 [-0.19 - 0.4] (p = 0.473) | 291 | 0.11 [-0.11 - 0.33] (p = 0.339) | p = 0.774 |
| Body Weight (kilograms) | 270 | 0.18 [-0.64 - 1.01] (p = 0.669) | 292 | 0.35 [-0.3 - 1.01] (p = 0.292) | p = 0.788 |
| Fat Mass (kilograms) | 269 | 0.14 [-0.24 - 0.52] (p = 0.473) | 291 | 0.22 [-0.09 - 0.52] (p = 0.164) | p = 0.537) |
| Fat percent (%) | 269 | 0.28 [-0.23 - 0.8] (p = 0.284) | 291 | 0.3 [-0.14 - 0.75] (p = 0.182) | p = 0.517 |
| Muscle Mass (kilograms) | 269 | 0.07 [-0.41 - 0.55] (p = 0.778) | 291 | 0.1 [-0.27 - 0.48] (p = 0.594) | p = 0.846 |
| Skeletal Mass (kilograms) | 269 | 0.04 [-0.25 - 0.33] (p = 0.776) | 291 | 0.06 [-0.16 - 0.28] (p = 0.59) | p = 0.844 |
| Bone Mass (kilograms) | 269 | 0 [-0.02 - 0.03] (p = 0.741) | 291 | 0 [-0.01 - 0.02] (p = 0.606) | p = 0.77 |
| Fat Free Mass (kilograms) | 269 | 0.07 [-0.44 - 0.58] (p = 0.776) | 291 | 0.11 [-0.29 - 0.5] (p = 0.594) | p = 0.843 |
| Fat Mass Index (FMI) | 269 | 0.08 [-0.09 - 0.24] (p = 0.362) | 291 | 0.09 [-0.04 - 0.22] (p = 0.168) | p = 0.551 |
| Fat-free Mass Index (FFMI) | 269 | 0.03 [-0.12 - 0.19] (p = 0.672) | 291 | 0.02 [-0.09 - 0.13] (p = 0.688) | p = 0.962 |
| **Body Anthropometrics COPSAC2000** | **n=** | **Female Adjusted [95% CI] p-value** | **n=** | **Male Adjusted [95% CI] p-value** | **P-Interaction** |
| BMI (actual) | 181 | 0.33 [-0.02 - 0.67] (p = 0.065) | 173 | 0.52 [0.28 - 0.76] (p < 0.001) | p = 0.328 |
| Body Weight (kilograms) | 181 | 0.8 [-0.2 - 1.79] (p = 0.12) | 175 | 1.72 [0.83 - 2.61] (p < 0.001) | p = 0.168 |
| Fat Mass (kilograms) | 181 | 0.52 [-0.11 - 1.15] (p = 0.106) | 173 | 1.07 [0.58 - 1.56] (p < 0.001) | p = 0.159 |
| Fat percent (%) | 181 | 0.37 [-0.12 - 0.86] (p = 0.145) | 173 | 0.84 [0.46 - 1.22] (p < 0.001) | p = 0.108 |
| Muscle Mass (kilograms) | 181 | 0.26 [-0.18 - 0.71] (p = 0.251) | 173 | 0.64 [0.15 - 1.12] (p = 0.011) | p = 0.287 |
| Skeletal Mass (kilograms) | 181 | 0.16 [-0.1 - 0.42] (p = 0.228) | 173 | 0.37 [0.07 - 0.67] (p = 0.015) | p = 0.335 |
| Bone Mass (kilograms) | 181 | 0.01 [-0.01 - 0.04] (p = 0.278) | 173 | 0.03 [0.01 - 0.06] (p = 0.007) | p = 0.255) |
| Fat Free Mass (kilograms) | 181 | 0.27 [-0.19 - 0.74] (p = 0.252) | 173 | 0.67 [0.16 - 1.18] (p = 0.011) | p = 0.285 |
| Fat Mass Index (FMI) | 181 | 0.2 [-0.02 - 0.43] (p = 0.082) | 173 | 0.33 [0.18 - 0.47] (p < 0.001) | p = 0.344 |
| Fat-free Mass Index (FFMI) | 181 | 0.12 [-0.02 - 0.27] (p = 0.091) | 173 | 0.19 [0.07 - 0.31] (p = 0.002) | p = 0.397 |

In COPSAC2000 at age 18 years, we found several significant associations between screen time and body anthropometrics, but the associations varied by sex. In females, screen time was associated with increased BMI (0.37 [0.02-0.72], p=0.039) and had borderline associations with increased body weight and fat mass. In contrast, male screen time was associated with all measured body anthropometrics, including BMI, body weight, fat mass, fat percentage, muscle mass, skeletal mass, bone mass, fat-free mass, fat mass index and fat-free mass index (p≤0.015) (*Table 3*), but with no significant interaction between sexes (p>0.13).

### Screen Time has a Distinct Blood Metabolome Signature and Associations with CVD Risk

We employed a machine learning prediction model in COPSAC2010, using screen time as the classifier and a targeted NMR blood metabolomics platform as the predictor. Our model considered 173 metabolic biomarkers and regularised to retain only the most influential predictors, resulting in a final model that included 37-screen time associated biomarkers (Figure S6). This model, with screen time z-scored for cross-dataset interpretation, trained in COPSAC2010 predicted screen time in both COPSAC2010 (0.80 [0.32-1.28], p=0.001), and COPSAC2000 (0.14 [0.03-0.26], p=0.014) (adjusted associations). These findings suggest that metabolic disturbances associated with screen time are a robust predictor of screen time across independent cohorts.

Additionally, we assessed the associations between screen time and cardiovascular risk in both COPSAC cohorts using an NMR Cardiovascular Risk Score, based on sex-stratified Cox proportional hazards models for 10-year CVD risk trained in the UK Biobank. Screen time showed a positive trend with the NMR CVD-score in COPSAC2010 at 10 years (0.06 [−0.02 - 0.13], p=0.15) and was significantly associated in COPSAC2000 at 18 years (0.07 [0.01 - 0.13], p=0.017) in adjusted models (*Table 2*).

### Sleep is a Modifying Factor in the Association between Screen Time and Cardiometabolic Risk

We explored how different lifestyle factors may influence the relationship between screen time and total CMR, focusing on lifestyle behaviours such as sedentary and light activity time, sleep duration and onset, and dietary patterns. Figure S7 visually represents the associations of these lifestyle factors across quartiles with screen time for COPSAC2010 and COPSAC2000, respectively. In interaction analysis, sedentary time, light activity time and a Western dietary pattern showed no significant contextual associations with screen time; however, sleep duration and onset emerged as modifiers in the screen time-CMR relationship.

In COPSAC2010, there was significant effect moderation between screen time and sleep duration (p=0.029), indicating that the positive association of screen time on CMR increased, as the amount of sleep decreased (*Figure 2A*). Similarly, there was significant effect moderation between screen time and sleep onset (p=0.009), suggesting that later sleep onset may exacerbate the detrimental effects of screen time on CMR (*Figure 2B*).

**Figure 2.**
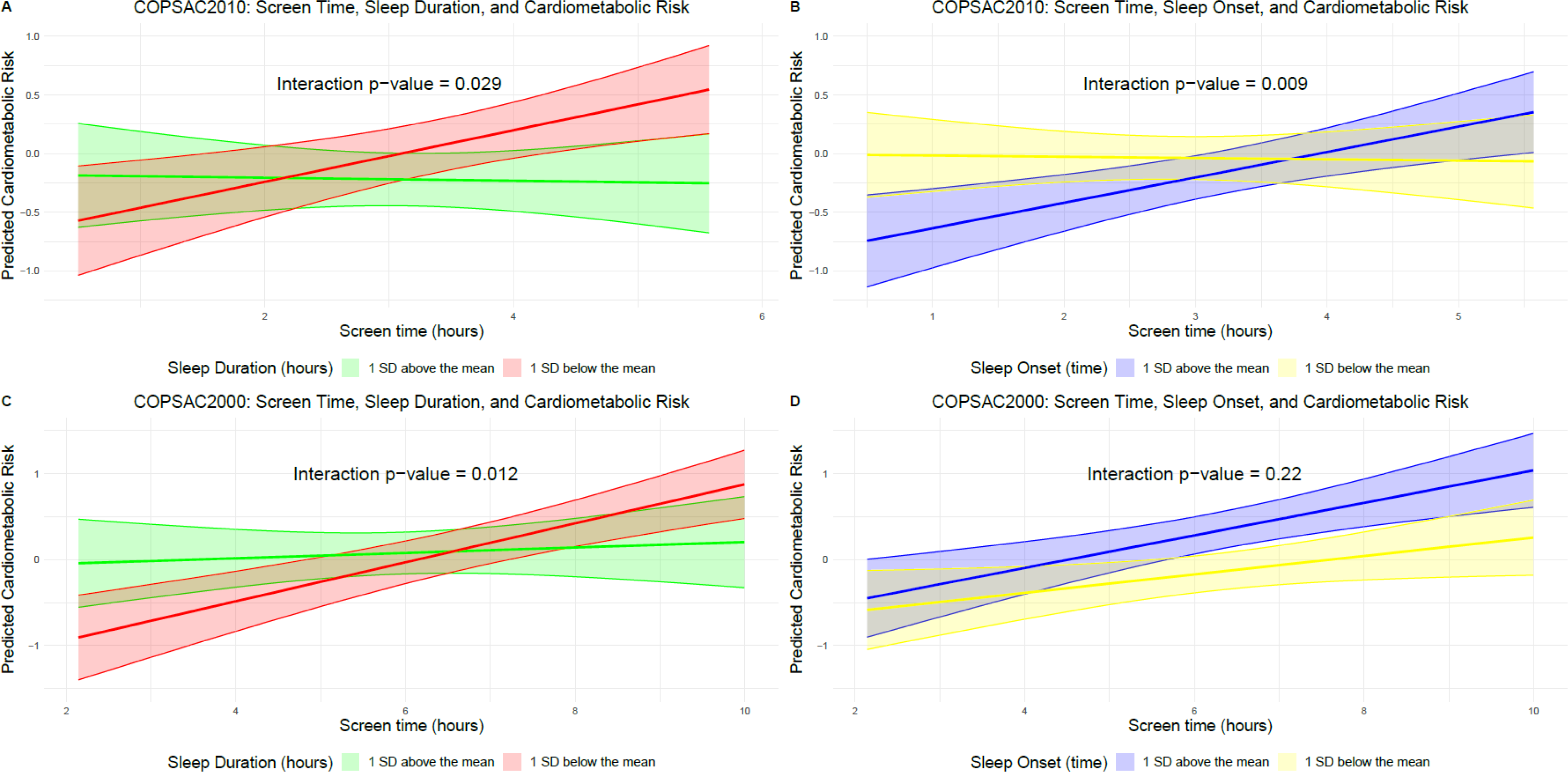
Modulating Effects of Sleep Duration and Onset on the Relationship between Screen Time and Cardiometabolic Risk in COPSAC2010 and COPSAC2000. Potential modulating effects of sleep duration and onset on the relationship between screen time and total cardiometabolic risk. **Panels A and B** depict these relationships for the COPSAC2010 cohort, while **Panels C and D** represent the COPSAC2000 cohort.

In COPSAC2000, there was a replication of this finding, with a significant moderation between screen time and sleep duration on total CMR (p=0.012) (*Figure 2C*). This suggests a similar trend as in COPSAC2010, whereby decreased sleep duration may exacerbate the negative impact of screen time on cardiometabolic health. There was a directional, but non-significant interaction between screen time and sleep onset time in COPSAC2000 (p=0.22) (*Figure 2D*).

We also conducted sex-stratified analyses to examine potential sex differences in moderating effects. In COPSAC2010 males, the interaction between screen time and sleep duration was more pronounced (p=0.031) (*Figure S8A*). In contrast, among COPSAC2000 females, the interaction between screen time and sleep onset was significant (p=0.005), suggesting that the detrimental moderation on CMR of increased screen time is magnified when the sleep onset time is later in female adolescents (*Figure S8B*).

## DISCUSSION

Our study establishes a clear association in two independent mother-child cohorts between screen time and elevated CMR in both childhood and adolescence, a relationship that persists even when accounting for objective measures of sleep, diet quality, physical activity and pubertal development. In childhood, lifestyle factors such as sleep duration and onset were found to significantly moderate the relationship between screen time and CMR, with sleep duration also partially mediating the association. In adolescence time of sleep onset had independent significant associations with CMR, and sleep duration significantly moderated the relationship. We employed a machine learning model trained in one cohort to uncover a unique metabolic signature associated with screen time, which despite significant cohort characteristic differences, was validated in the other cohort thus opening an avenue for identifying individuals at particular risk of high screen time use. Finally, we used blood metabolome profiles to assess cardiovascular risk using adult data trained on actual CVD outcomes, finding positive directional associations with screen time in childhood and significant associations in adolescence.

Previous studies have reported no association between self-reported screen time and CMR in children aged 7-12 (18). However, our study indicates that such an association becomes more pronounced in adolescence, suggesting that the impact of screen time on cardiometabolic health may evolve throughout childhood and adolescence. This is supported by a further study which found that higher screen time in adolescence was associated with higher odds of select indicators of cardiometabolic disease in adulthood, including obesity, hypertension, hyperlipidemia, and diabetes (6). Our study further enriches the literature by identifying sleep duration and time of onset as key modulating factors in the screen time-CMR association in childhood. We validated the moderating effect of sleep duration on CMR in COPSAC2000 and found that time of sleep onset was a significant independent risk factor in adolescence. However, a recent study of 3000 participants aged 6-17 utilising self-reported measures of sleep and screen time, found no moderating role of sleep on screen times associated with cardiometabolic risk (31). Our findings align with existing literature that link shorter sleep duration with higher CMR in adolescents (32).

A unique strength of our study is the use of two prospective mother-child cohorts with deep phenotyping and longitudinal follow-up. The COPSAC2010 cohort, in particular, utilised repeated outcome and exposure data, allowing for us to account for inter-individual variation in participants, thus providing more robust inference. Furthermore, our application of a machine learning approach uncovered a robust metabolic signature of screen time, which was a significant predictor of actual screen time, independent of other potential confounding factors. To the best of our knowledge, we are also the first study to implement CVD-risk scores modelled on adult CVD outcomes in independent childhood and implement these for inference. Thus, our comprehensive approach which accounts for objective lifestyle measures, establishes a robust link between screen time and CMR during both childhood and adolescence. However, our study also has limitations. The observational nature of the study design limits our ability to infer causality, and we cannot rule out residual confounding. Furthermore, our reliance on reported measures for screen time could introduce bias (13). However, this methodology is widely accepted in the field (33), and the prevalence of high screen time usage is corroborated by studies using wearable cameras to monitor children’s screen activities (34). Furthermore, while many of our lifestyle measures such as sleep and physical activity were objective, our dietary assessments were self-reported and therefore potentially subject to recall or social desirability bias. Finally, the nature of our moderation modelling of lifestyle factors and mediation analysis were exploratory, and constituted subanalysis’. Therefore, while these findings provide valuable context, they should be interpreted with caution and considered as hypothesis-generating, rather than definitive.

Moving forward, future studies should focus on using objective measures of screen time and related lifestyle behaviours. Our findings suggest that sleep duration and onset may play a significant role in moderating the impact of screen time on cardiometabolic health in childhood. Associations between sleep duration and cardiometabolic risk in childhood and adolescence are well documented (35), with suggested mechanisms including circadian rhythm misalignment (36), sodium retention secondary to insulin resistance (37) and increased sympathetic nervous system activity (35). Our findings indicate a contextual association between screen time and cardiometabolic risk, which becomes stronger in cases of decreased sleep. This may be explained by the reduction in melatonin levels caused by exposure to screen light in the evening, leading to disturbances in circadian rhythms that exacerbate this relationship (38). Moreover, our study suggests that the associations between screen time and cardiometabolic risk are independent of lifestyle behaviours, including sedentary time, suggesting that independent mechanisms whereby screen time may influence cardiometabolic risk, such as poor stress regulation and high sympathetic arousal (39).

Future studies are needed to confirm and further elucidate these relationships. Our results also highlight the potential utility of machine learning approaches in identifying key metabolic signatures of screen time. Despite the parental-/self-reported nature of our screen time data, our model was able to predict screen time across two distinct cohorts with significantly different baseline characteristics, suggesting that the metabolic signature of screen time is robust and consistent across varying populations. These insights could be useful in future research to both identify individuals at risk of high screen time and its associated CMR and inform interventions to reduce screen time and improve cardiometabolic health.

In conclusion, our findings from two independent mother-child cohorts emphasise the significant detrimental impact of screen time on cardiometabolic risk, with sleep duration and time of onset acting as key contextual factors. Distinct patterns of associations when juxtaposing these cohorts suggests that interventions aimed at reducing CMR may need to be tailored distinctly for children and adolescents, considering the varying influences of lifestyle factors at these stages. These insights underline the importance of comprehensive, multifaceted strategies to mitigate CMR in childhood and adolescence, with a particular focus on reducing screen time, and promoting healthier sleeping habits.

## Data Availability

Participant-level personally identifiable data are protected under the Danish Data Protection Act and European Regulation 2016/679 of the European Parliament and of the Council (GDPR) that prohibit distribution even in pseudo-anonymized form. However, participant-level data can be made available under a data transfer agreement as part of a collaboration effort.

## Abbreviations

ADHD: Attention-deficit/hyperactivity disorder
ADHD-RS: ADHD Rating Scale
ApoB: Apolipoprotein B
ASRS: Adult ADHD Self-Report Scale
BIA: Bioelectrical impedance analysis
BMI: Body mass index
CMR: Cardiometabolic risk
COPSAC2000: COpenhagen Prospective Studies on Asthma in Childhood mother-child cohort 2000
COPSAC2010: COpenhagen Prospective Studies on Asthma in Childhood mother-child cohort 2010
CVD: Cardiovascular disease
FFM: Fat-free mass
FMI: Fat mass index
FFMI: Fat-free mass index
FSH: Follicle stimulating hormone
HDL: High-density lipoprotein
HOMA-IR: Homeostatic Model Assessment for Insulin Resistance
hs-CRP: High-sensitivity CRP
LH: Luteinizing hormone
n3-LCPUFA: Omega-3 long-chain polyunsaturated fatty acids
SBP: Systolic Blood Pressure
sPLS: Sparse partial least square

